# Long-term clinical outcomes of bariatric surgery in adults with severe obesity: A population-based retrospective cohort study

**DOI:** 10.1101/2023.05.31.23290770

**Authors:** Natasha Wiebe, Marcello Tonelli

## Abstract

**Background:** Bariatric surgery leads to sustained weight loss in a majority of recipients, and also reduces fasting insulin levels and markers of inflammation. We described the long-term associations between bariatric surgery and clinical outcomes including 30 morbidities.

**Methods:** We did a retrospective population-based cohort study of 304,157 adults with severe obesity, living in Alberta, Canada; 6,212 of whom had bariatric surgery. We modelled adjusted time to mortality, hospitalization, surgery and the adjusted incidence/prevalence of 30 morbidities after 5 years of follow-up.

**Results:** Over a median follow-up of 4.4 years (range 1 day-22.0 years), bariatric surgery was associated with increased risk of hospitalization (HR 1.46, 95% CI 1.41,1.51) and additional surgery (HR 1.42, 95% CI 1.32,1.52) but with a decreased risk of mortality (HR 0.76, 95% CI 0.64,0.91). After 5 years, bariatric surgery was associated with a lower risk of severe chronic kidney disease (HR 0.45, 95% CI 0.27,0.75), coronary disease (HR 0.49, 95% CI 0.33,0.72), diabetes (HR 0.51, 95% CI 0.47,0.56), inflammatory bowel disease (HR 0.55, 95% CI 0.37,0.83), hypertension (HR 0.70, 95% CI 0.66,0.75), chronic pulmonary disease (HR 0.75, 95% CI 0.66,0.86), asthma (HR 0.79, 95% 0.65,0.96), cancer (HR 0.79, 95% CI 0.65,0.96), and chronic heart failure (HR 0.79, 95% CI 0.64,0.96). In contrast, after 5 years, bariatric surgery was associated with an increased risk of peptic ulcer (HR 1.99, 95% CI 1.32,3.01), alcohol misuse (HR 1.55, 95% CI 1.25,1.94), frailty (HR 1.28, 95% 1.11,1.46), severe constipation (HR 1.26, 95% CI 1.07,1.49), sleep disturbance (HR 1.21, 95% CI 1.08,1.35), depression (HR 1.18, 95% CI 1.10,1.27), and chronic pain (HR 1.12, 95% CI 1.04,1.20).

**Interpretation:** Bariatric surgery was associated with lower risks of death and certain morbidities. However, bariatric surgery was also associated with increased risk of hospitalization and additional surgery, as well as certain other morbidities. Since values and preferences for these various benefits and harms may differ between individuals, this suggests that comprehensive counselling should be offered to patients considering bariatric surgery.

## Introduction

Bariatric surgery leads to sustained weight loss in most recipients, and also reduces fasting insulin levels^1, 2^ and markers of inflammation.^3^ Bariatric surgery can reverse and/or lessen the severity of diabetes and hypertension,^4^ and likely obstructive sleep apnea.^5, 6^ However, bariatric surgery is also associated with an excess risk of certain adverse outcomes, including venous thrombosis, reactive hypoglycemia,^7^ nephrolithiasis,^8^ fractures,^9^ depression, alcohol misuse, and suicide.^10^

The long-term risks and benefits of bariatric surgery have been incompletely studied. Few studies examine the frequency of additional surgery and hospitalizations following the index bariatric procedure, and most have considered a relatively narrow range of adverse clinical outcomes. The current paradigm is that the benefits of bariatric surgery are mediated by reductions in fat mass. However, higher BMI and fat percentage do not correlate with excess mortality after adjustment for fasting insulin and chronic inflammation in the general population, and in fact appear paradoxically protective.^11, 12^ Hyperinsulinemia and chronic inflammation have also been linked to a wide range of health conditions, including cardiovascular disease, diabetes, fatty liver, sleep apnea, cancer, chronic obstructive pulmonary disorder, dementia, and autoimmune disorders.^13, 14^ Given the beneficial effects of bariatric surgery on hyperinsulinemia as well as inflammatory biomarkers, it is possible that these effects contribute to the clinical benefits of bariatric surgery, which in turn may extend beyond those conditions that have been studied to date. Equally, given that bariatric surgery does not always produce weight loss (failure ranges from 11% to 35% in Roux-en-Y gastric bypass^15^) and can cause complications from anatomical abnormalities of the gastrointestinal tract (e.g., malabsorption, nutritional deficiency, bacterial overgrowth) which can themselves lead to other health conditions (e.g., osteoporosis, peripheral neuropathy, depression), a comprehensive assessment of clinical outcomes is warranted.

In this retrospective population-based cohort study of adults with severe obesity, we aimed to describe the long-term effects of bariatric surgery on clinical outcomes such as mortality, hospitalization, and additional surgery, as well as the risk of 30 morbidities during follow-up.

## Methods

We report this retrospective population-based cohort study according to the STROBE guidelines.^16^ The institutional review boards at the Universities of Alberta (Pro00053469) and Calgary (REB16-1575) approved this study and waived the requirement for participants to provide consent due to the large sample size. We did analyses between December 2022 and April 2023.

### Data sources and cohort

We used the Alberta Kidney Disease Network database,^17–19^ which incorporates data from Alberta Health (AH; the provincial health ministry) including data on registration, vital statistics, provider claims, hospitalizations and ambulatory care utilization; and from the clinical laboratories in Alberta. All adults registered with AH were included in the database; all Alberta residents are eligible for insurance coverage by AH and >99% participate in coverage. We used the database to assemble a cohort of adults with severe obesity who resided in Alberta, Canada. We followed participants until death, out-migration or study end (March 31, 2019), whichever was earliest.

### Bariatric surgery and severe obesity

We determined the incidence of bariatric surgery using procedure codes from provider claims (ICD-9 CCP codes; Supplemental eTable 1). The date of the first procedure code was considered the index date for participants with bariatric surgery (cases); these dates for bariatric surgery could range from April 1, 1997 to December 31, 2018, ensuring at least 90 days of follow-up after surgery. After matching on entry date (90 days within registration with AH, 18^th^ birthday or study start, whichever date was latest), the index dates for participants without bariatric surgery (controls) were randomly drawn with replacement from the distribution of bariatric surgery dates in cases. We excluded control participants with index dates after their end dates from the cohort.

We identified the specific type of bariatric surgery using the following claim codes: Roux-en-Y gastric bypass (56.93A), sleeve gastrectomy (56.93C), adjustable gastric band (56.93B, 56.93F), and unknown (56.93). In a sensitivity analysis, we used a definition from the Canadian Institute for Health Information (CIHI; www.cihi.ca), which identifies bariatric surgery, using hospitalization with ICD-10-CA CCI codes (1.NF.78.^^) which were converted to ICD-9-CM procedure codes (43.89, 44.39).^20^ This latter definition was similar to previous publications in bariatric surgery using administrative data.^21–24^ We excluded bariatric codes if a participant was hospitalized, at the same time, for a gastrointestinal or abdominal cancer, or a perforated gastrointestinal ulcer (Supplemental eTable 1).

We considered severe obesity to be present if there was a procedure marked with a BMI supplemental fee modifier as in our prior work^25^ (≥35 kg/m^2^ before 2017 and ≥40 kg/m^2^ in/after 2017; Supplemental eTable 1). From our parent dataset of registered adult Albertans, 6.1% had severe obesity and thus formed the cohort for this study (Figure 1).

**Figure 1.**
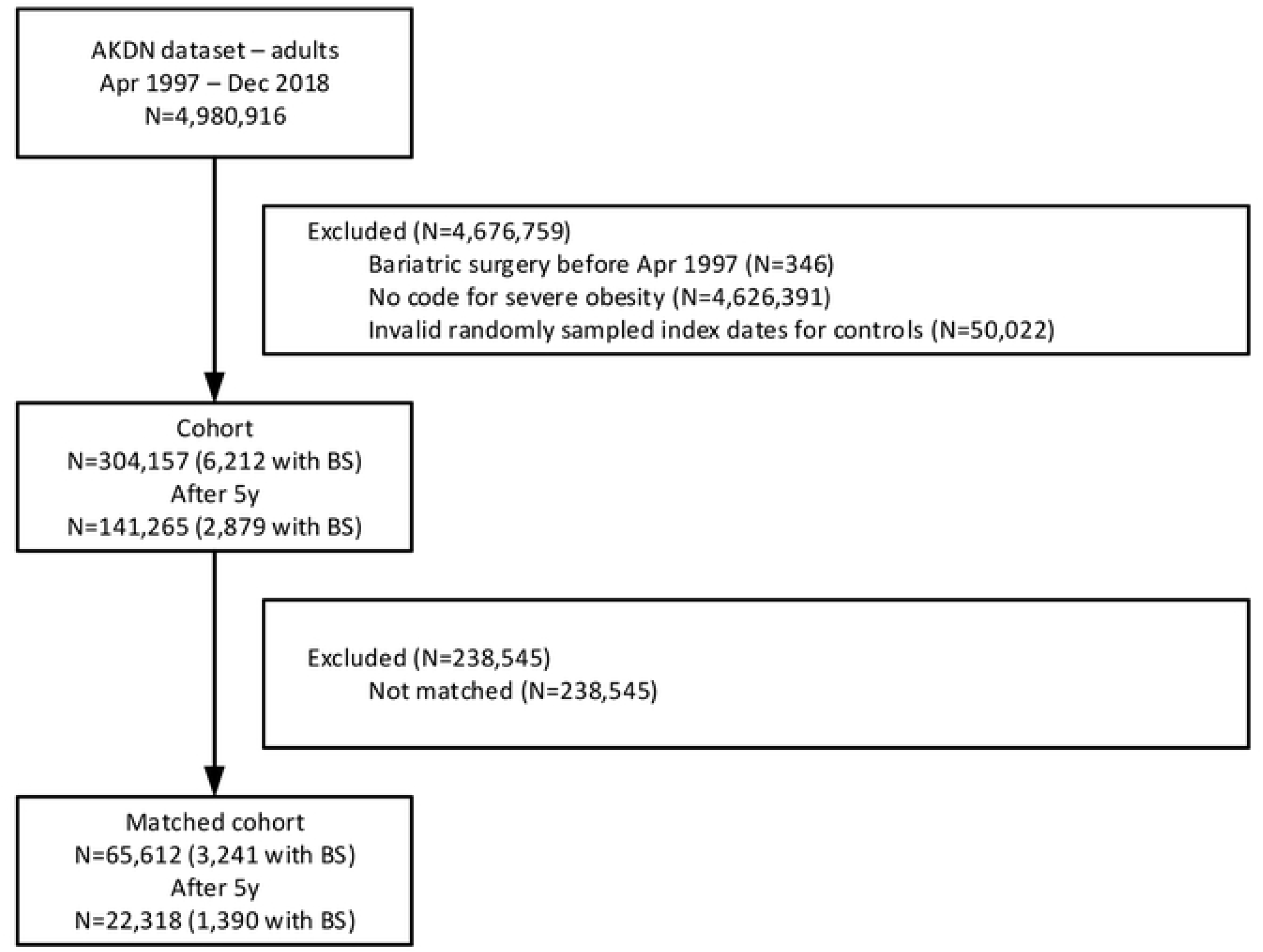
Participant flow diagram. AKDN Alberta Kidney Disease Network, BS bariatric surgery Randomly sampled index dates for controls were invalid if the index date occurred after their end date.

### Morbidities and other covariates

We defined morbidities using a previously published framework with validated algorithms as applied to Canadian provider claims data, each of which had positive predictive values ≥70% as compared to a gold standard measure such as chart review.^26^ Morbidities include those potentially related to hyperinsulinemia (e.g., cardiovascular disease), inflammation (e.g., respiratory diseases), nutritional deficiencies (e.g., fragility fractures), anatomical modifications of the gastrointestinal system (e.g., peptic ulcer disease), combinations of the above, or none of the above (e.g., multiple sclerosis): alcohol misuse, asthma, atrial fibrillation, cancer (non-metastatic breast, cervical, colorectal, pulmonary, and prostate cancers, metastatic cancer, and lymphoma), chronic heart failure, chronic pain, chronic pulmonary disease, coronary artery disease (CAD; myocardial infarction, percutaneous intervention, coronary artery bypass graft), dementia, depression, diabetes, epilepsy, frailty^27^ (fragility fractures: wrist, forearm, humerus, hip, pelvis, and spine; and/or osteoporosis), gout,^28^ hypertension, hypothyroidism, inflammatory bowel disease (IBD), irritable bowel syndrome, liver disease (chronic hepatitis B, cirrhosis), multiple sclerosis, Parkinson’s disease, peptic ulcer disease (PUD), peripheral artery disease, psoriasis, rheumatic disease, schizophrenia, severe constipation, sleep disturbance,^29^ and stroke or transient ischemic attack (TIA). Detailed methods for classifying comorbidity status and the specific algorithms used are found elsewhere.^26^ Our dataset did not permit us to identify obstructive sleep apnea (OSA) specifically, and so we modified the previously validated algorithm to include any sleep disturbance (2 claims within 2 years: ICD-9 327, 347, and 780.5).^29^ We also considered severe chronic kidney disease (CKD) as a 30^th^ condition, which was defined by a sustained estimated glomerular filtration rate below 30 mL/min per 1.73m^2^. We classified each participant with respect to the presence or absence of these 30 chronic conditions at baseline (lookback extended as far as April 1994 where records were available)^30^ and after a 5-year washout period following surgery to allow morbidities to reverse after bariatric surgery.

As in our prior work we used administrative data to identify age, biological sex, and rural residence location.^31^ We included the Pampalon index of material deprivation,^32, 33^ which categorizes participants based on their residential postal code into 5 bins of socioeconomic inequalities in health care services and population health with 5 representing the most deprived neighbourhoods. Rural status (1.8%) and material deprivation (3.0%) were missing a few values but no other variables had missing values.

### Outcomes

Clinical outcomes were all-cause death, hospitalization, surgery (in the first 90 days, in the first 5 years [not including the index bariatric surgery], after 5 years, and until end of follow-up), and exclusively after 5 years (a washout period), we considered new (incident) or ongoing cases of the 30 morbidities listed earlier. Participants with dementia, multiple sclerosis, Parkinson’s disease, and schizophrenia at baseline were not included in analyses evaluating the incidence of these outcomes.

### Statistical analyses

We did analyses with Stata MP 17·0 (www.stata.com) and reported baseline descriptive statistics as counts and percentages, or means and standard deviations, as appropriate. Differences between cases and controls were tested using logistic regression.

We used unadjusted, age-adjusted and fully-adjusted Weibull hazards regression to determine the associations between bariatric surgery and time to death, hospitalization, surgery, and continuation/incidence of each comorbidity. In the fully-adjusted models, we regressed outcomes on age, biological sex, rural residence, neighbourhood material deprivation index, date of first obesity modifier, and 30 morbidities. We determined that the proportional hazard assumption was satisfied by examining plots of the log-negative-log of within-group survivorship probabilities versus log-time after adjustment for covariates. We report hazard ratios with 95% confidence intervals (in text and with forest plots) and set the threshold *p* for statistical significance at 0.05.

We did several sensitivity analyses. One, in an effort to achieve strong covariate balance, we curated a cohort with (many to many) exact matches for every demographic characteristic, obesity year, and morbidity; we grouped age into 5-year bins. We opted not to include inverse- weight propensity scoring analyses as they can increase imbalance between intervention groups.^34^ Two, in the after-5-year outcomes, we modelled death as a competing risk using a flexible modelling approach outlined by Lambert.^35^ We retained participants who had died in the first 5 years in this cohort; their outcome data were imputed and weighted (using the Stata command *stcrprep*) based on the conditional probability of being censored with or without bariatric surgery. Three, we regressed the number of hospitalizations, the number of days in hospital, and the number of surgeries onto bariatric surgery and all covariates using negative binomial models. Four, we parameterized follow-up duration using restricted cubic splines using 4 knots (at 1, 5, 10, and 15 years) and 6 knots (at 90 days, 1, 5, 10, 15, and 20 years). We regressed time to death on the main effects and interactions of bariatric surgery with the splines of follow-up, and further adjusted for all covariates. Five, we interacted fiscal year of the index date (as a linear term) with bariatric surgery and reported estimates for the 2000, 2005, 2010, and 2015 fiscal years. We based the 2015 estimates on extrapolations for the after-5-year outcomes. All sensitivity analyses were fully adjusted.

In order to illustrate the number of adverse outcomes averted and experienced, we calculated the numbers needed to benefit and harm using an approach by Altman and Andersen.^36^ These estimates were inverted and multiplied by 400 in order to depict the numbers of outcomes averted or experienced in a hypothetical population of 400 recipients of bariatric surgery.

## Results

### Characteristics of study participants

Participant flow is shown in Figure 1. Overall most 4,626,391 (93.8%) adult Albertans were excluded because they did not meet our definition of severe obesity. Of those with severe obesity, 6,212 (2.0%) had undergone bariatric surgery and 297,945 (98.0%) had not. The quantile-quantile plot shows that the distribution of index dates was similar for cases and controls (Supplemental eFigure 1). The 304,157 participants were followed for a median of 4.4 years (range 1 day to 22.0 years). There were 15,670 deaths (5.2%); 140,121 (46.1%) participants had at least one hospitalization (for a total of 363,744 hospitalizations), and 24,165 (7.9%) had at least one additional surgery (for a total of 29,062 surgeries after the index bariatric surgery) during follow-up.

Table 1 summarizes demographics and clinical characteristics by receipt of bariatric surgery or not. Participants who had undergone bariatric surgery were younger, more likely to be female, and to reside in an urban residence. The morbidities were sorted by the likelihood of receiving bariatric surgery, from most to least, after adjustment for all covariates. In order of magnitude, participants were more likely to receive bariatric surgery if they had sleep disturbance, hypertension, PUD, asthma, diabetes, depression, psoriasis, hypothyroidism, chronic pain, and gout. Also, in order of magnitude, participants were less likely to receive bariatric surgery if they had severe CKD, alcohol misuse, CAD, liver disease, schizophrenia, Parkinson’s disease, dementia, cancer, heart failure, severe constipation, IBD, stroke/TIA, epilepsy, frailty, and rheumatic disease. While participants who had bariatric surgery had more morbidities on average (mean of 2.3 versus 1.8), they were more likely to have morbidities with relatively better prognosis (e.g., hypertension, depression, chronic pain) and participants without bariatric surgery were more likely to have conditions that have relatively worse prognosis (e.g., heart failure, liver disease, severe CKD). Furthermore, in the 5 years prior to the index date, the participants without bariatric surgery had more hospitalizations and spent more days in hospital.

**Table 1.** Demographic and clinical characteristics by bariatric surgery.

| Characteristics | Full cohort |  | P | Exact matches |  |
| --- | --- | --- | --- | --- | --- |
|  | Bariatric | No bariatric |  | Bariatric | No bariatric |
| N | 6,212 | 297,945 | - | 3,241 | 62,371 |
| Age, y | 43.7 [10.5] | 48.3 [16.1] | <0.001 | 41.6 [10.2] | 41.6 [10.3] |
| Male | 918 (14.8) | 90,034 (30.2) | <0.001 | 364 (11.2) | 7,005 (11.2) |
| Rural residence | 660 (10.7) | 43,163 (14.8) | <0.001 | 221 (6.9) | 4,253 (6.9) |
| Material deprivation | 3.3 [1.4] | 3.3 [1.4] | 0.71 | 3.3 [1.3] | 3.3 [1.3] |
| Morbidities | 2.3 [1.7] | 1.8 [2.0] | <0.001 | 1.3 [1.0] | 1.3 [1.0] |
| Sleep disturbance | 843 (13.6) | 8,866 (3.0) | <0.001 | 72 (2.2) | 1,386 (2.2) |
| Hypertension | 2,960 (47.6) | 115,025 (38.6) | <0.001 | 1,154 (35.6) | 22,208 (35.6) |
| Peptic ulcer disease | 19 (0.3) | 431 (0.1) | 0.001 | 0 (0.0) | 0 (0.0) |
| Asthma | 827 (13.3) | 15,131 (5.1) | <0.001 | 141 (4.4) | 2,713 (4.4) |
| Diabetes | 1,828 (29.4) | 56,230 (18.9) | <0.001 | 582 (18.0) | 11,200 (18.0) |
| Depression | 1,995 (32.1) | 43,538 (14.6) | <0.001 | 738 (22.8) | 14,202 (22.8) |
| Psoriasis | 94 (1.5) | 2,893 (1.0) | <0.001 | 5 (0.2) | 96 (0.2) |
| Hypothyroid | 1,055 (17.0) | 35,903 (12.1) | <0.001 | 339 (10.5) | 6,524 (10.5) |
| Chronic pain | 1,637 (26.4) | 56,706 (19.0) | <0.001 | 603 (18.6) | 11,604 (18.6) |
| Gout | 445 (7.2) | 23,979 (8.0) | 0.01 | 85 (2.6) | 1,636 (2.6) |
| IBS | 239 (3.8) | 8,238 (2.8) | <0.001 | 26 (0.8) | 500 (0.8) |
| Chronic pulmonary | 812 (13.1) | 38,094 (12.8) | 0.50 | 164 (5.1) | 3,156 (5.1) |
| Multiple sclerosis | 60 (1.0) | 2,639 (0.9) | 0.51 | 4 (0.1) | 77 (0.1) |
| PAD | 41 (0.7) | 3,350 (1.1) | <0.001 | 1 (0.0) | 19 (0.0) |
| Atrial fibrillation | 134 (2.2) | 11,733 (3.9) | <0.001 | 8 (0.2) | 154 (0.2) |
| Rheumatic disease | 136 (2.2) | 7,025 (2.4) | 0.39 | 3 (0.1) | 58 (0.1) |
| Frailty | 479 (7.7) | 28,811 (9.7) | <0.001 | 76 (2.3) | 1,463 (2.3) |
| Epilepsy | 105 (1.7) | 5,592 (1.9) | 0.28 | 2 (0.1) | 38 (0.1) |
| Stroke/TIA | 208 (3.3) | 16,148 (5.4) | <0.001 | 18 (0.6) | 346 (0.6) |
| IBD | 67 (1.1) | 4,122 (1.4) | 0.04 | 4 (0.1) | 77 (0.1) |
| Severe constipation | 61 (1.0) | 3,762 (1.3) | 0.05 | 3 (0.1) | 58 (0.1) |
| Chronic heart failure | 146 (2.4) | 13,950 (4.7) | <0.001 | 3 (0.1) | 58 (0.1) |
|  | Bariatric | No bariatric |  | Bariatric | No bariatric |
| Cancer | 98 (1.6) | 11,323 (3.8) | <0.001 | 15 (0.5) | 289 (0.5) |
| Dementia | 27 (0.4) | 3,637 (1.2) | <0.001 | 0 (0.0) | 0 (0.0) |
| Parkinson's | 10 (0.2) | 1,269 (0.4) | 0.002 | 0 (0.0) | 0 (0.0) |
| Schizophrenia | 64 (1.0) | 3,977 (1.3) | 0.04 | 3 (0.1) | 58 (0.1) |
| Liver disease | 8 (0.1) | 1,084 (0.4) | 0.003 | 0 (0.0) | 0 (0.0) |
| CAD | 65 (1.0) | 11,004 (3.7) | <0.001 | 3 (0.1) | 58 (0.1) |
| Alcohol misuse | 81 (1.3) | 9,113 (3.1) | <0.001 | 7 (0.2) | 135 (0.2) |
| Severe CKD | 7 (0.1) | 1,698 (0.6) | <0.001 | 0 (0.0) | 0 (0.0) |
| In the prior 5y |  |  |  |  |  |
| Hospitalizations | 0.6 [1.3] | 0.8 [1.6] | <0.001 | 0.5 [0.8] | 0.6 [1.0] |
| LOS, d | 3.1 [16.4] | 5.6 [30.2] | <0.001 | 1.5 [6.9] | 1.9 [6.7] |

CAD coronary artery disease, CKD chronic kidney disease, IBD inflammatory bowel disease, IBS irritable bowel syndrome, LOS length of stay, PAD peripheral artery disease, SD standard deviation, TIA transient ischemic attack
N (%) or mean (SD) as appropriate. Morbidities are sorted by the adjusted odds of receiving bariatric surgery (the adjusted odds ratios are not shown).

### All-cause mortality

In unadjusted analyses, bariatric surgery was associated with significantly lower mortality before and after 5 years of follow-up (HR 0.24, 95% CI 0.17,0.33, and HR 0.48, 95% CI 0.39,0.60, respectively; Table 2). With full adjustment, the risk associated with bariatric surgery remained significantly lower before 5 years of follow-up (HR 0.58, 95% CI 0.42,0.81; Figure 2), but not after 5 years of follow-up (HR 0.95, 95% CI 0.77,1.19). Over all of follow-up, the adjusted risk for mortality was significantly lower (HR 0.76, 95% CI 0.64,0.91).

**Table 2.** Time-to-event outcomes associated with bariatric surgery.

| **Outcomes** | **N** | **Events (%)** | **Unadjusted** | **Age-adjusted** | **Fully adjusted** |
| --- | --- | --- | --- | --- | --- |
| **Mortality** |  |  |  |  |  |
| All of follow-up | 304,157 | 15,670 (5.2) | **0.37 (0.31,0.44)** | 0.92 (0.77,1.10) | **0.76 (0.64,0.91)** |
| First 90 days | 304,157 | 490 (0.2) | 0.80 (0.40,1.60) | 1.97 (0.97,4.00) | 1.91 (0.93,3.89) |
| First 5 years | 304,157 | 7,410 (2.4) | **0.24 (0.17,0.33)** | **0.68 (0.49,0.94)** | **0.58 (0.42,0.81)** |
| After 5 years | 141,265 | 8,260 (5.9) | **0.48 (0.39,0.60)** | 1.20 (0.97,1.50) | 0.95 (0.77,1.19) |
| **Hospitalization** |  |  |  |  |  |
| All of follow-up | 304,157 | 140,121 (46.1) | **1.58 (1.53,1.64)** | **1.64 (1.59,1.70)** | **1.46 (1.41,1.51)** |
| First 90 days | 304,157 | 12,041 (4.0) | **2.71 (2.50,2.94)** | **2.92 (2.69,3.17)** | **2.58 (2.37,2.81)** |
| First 5 years | 304,157 | 104,714 (34.4) | **1.68 (1.62,1.74)** | **1.74 (1.68,1.80)** | **1.54 (1.49,1.60)** |
| After 5 years | 141,265 | 65,329 (46.3) | **1.20 (1.14,1.26)** | **1.31 (1.24,1.38)** | **1.09 (1.03,1.15)** |
| **Surgery** |  |  |  |  |  |
| All of follow-up | 304,157 | 24,165 (7.9) | **1.79 (1.67,1.92)** | **1.61 (1.50,1.73)** | **1.42 (1.32,1.52)** |
| First 90 days | 304,157 | 1,273 (0.4) | **2.89 (2.28,3.66)** | **2.68 (2.11,3.40)** | **2.23 (1.74,2.85)** |
| First 5 years | 304,157 | 15,826 (5.2) | **1.76 (1.62,1.92)** | **1.61 (1.48,1.76)** | **1.44 (1.32,1.57)** |
| After 5 years | 141,265 | 9,781 (6.9) | **1.87 (1.68,2.08)** | **1.63 (1.47,1.82)** | **1.38 (1.24,1.54)** |
| **After 5 years** |  |  |  |  |  |
| Severe CKD | 141,265 | 2,940 (2.1) | **0.24 (0.15,0.41)** | 0.67 (0.40,1.12) | **0.45 (0.27,0.75)** |
| CAD | 141,265 | 4,615 (3.3) | **0.26 (0.17,0.38)** | **0.40 (0.27,0.59)** | **0.49 (0.33,0.72)** |
| Diabetes | 141,265 | 37,136 (26.3) | **0.71 (0.65,0.77)** | 0.95 (0.88,1.04) | **0.51 (0.47,0.56)** |
| IBD | 141,265 | 1,750 (1.2) | 0.69 (0.47,1.03) | 0.71 (0.48,1.05) | **0.55 (0.37,0.83)** |
| Hypertension | 141,265 | 63,825 (45.2) | **0.68 (0.64,0.73)** | 0.98 (0.92,1.04) | **0.70 (0.66,0.75)** |
| PAD | 141,265 | 2,394 (1.7) | **0.46 (0.31,0.70)** | 0.85 (0.57,1.29) | 0.77 (0.51,1.17) |
| Chronic pulmonary | 141,265 | 16,085 (11.4) | **0.67 (0.59,0.76)** | 1.03 (0.90,1.18) | **0.75 (0.66,0.86)** |
| Asthma | 141,265 | 3,096 (2.2) | **1.75 (1.44,2.12)** | **1.82 (1.50,2.21)** | **0.79 (0.65,0.96)** |
| Cancer | 141,265 | 9,499 (6.7) | **0.54 (0.44,0.65)** | **0.79 (0.66,0.96)** | **0.79 (0.65,0.96)** |
| Chronic heart failure | 141,265 | 10,841 (7.7) | **0.44 (0.36,0.54)** | 1.01 (0.83,1.24) | **0.79 (0.64,0.96)** |

| Outcomes | N | Events (%) | Unadjusted | Age-adjusted | Fully adjusted |
| --- | --- | --- | --- | --- | --- |
| Liver disease | 141,265 | 1,166 (0.8) | **0.58 (0.34,0.99)** | 0.77 (0.46,1.31) | 0.79 (0.46,1.35) |
| Atrial fibrillation | 141,265 | 8,856 (6.3) | **0.43 (0.35,0.54)** | 0.92 (0.74,1.15) | 0.84 (0.67,1.04) |
| Gout | 141,265 | 10,725 (7.6) | **0.62 (0.53,0.73)** | **0.84 (0.71,0.996)** | 0.86 (0.73,1.02) |
| Psoriasis | 141,265 | 987 (0.7) | 1.25 (0.84,1.86) | 1.33 (0.89,1.98) | 0.87 (0.57,1.31) |
| Stroke/TIA | 141,265 | 7,817 (5.5) | **0.63 (0.52,0.77)** | 1.04 (0.85,1.26) | 0.88 (0.72,1.07) |
| IBS | 141,265 | 1,878 (1.3) | **1.52 (1.16,1.97)** | **1.48 (1.14,1.93)** | 0.88 (0.67,1.15) |
| Schizophrenia | 139,752 | 693 (0.5) | **1.57 (1.03,2.40)** | **1.55 (1.01,2.37)** | 0.88 (0.57,1.36) |
| Multiple sclerosis | 140,261 | 695 (0.5) | 1.27 (0.80,2.04) | **1.26 (0.79,2.02)** | 1.00 (0.62,1.62) |
| Hypothyroid | 141,265 | 16,183 (11.5) | **1.33 (1.21,1.47)** | **1.53 (1.39,1.69)** | 1.01 (0.91,1.11) |
| Rheumatic disease | 141,265 | 16,183 (11.5) | **1.33 (1.21,1.47)** | **1.53 (1.39,1.69)** | 1.01 (0.91,1.11) |
| Dementia | 140,735 | 3,619 (2.6) | **0.44 (0.31,0.62)** | **1.45 (1.03,2.05)** | 1.06 (0.75,1.51) |
| Parkinson's | 140,903 | 775 (0.6) | 0.69 (0.38,1.25) | 1.37 (0.75,2.49) | 1.10 (0.60,2.01) |
| Chronic pain | 141,265 | 33,966 (24.0) | **1.32 (1.23,1.41)** | **1.40 (1.30,1.50)** | **1.12 (1.04,1.20)** |
| Depression | 141,265 | 25,765 (18.2) | **1.81 (1.69,1.94)** | **1.77 (1.65,1.89)** | **1.18 (1.10,1.27)** |
| Sleep disturbance | 141,265 | 10,896 (7.7) | **1.49 (1.34,1.67)** | **1.67 (1.49,1.87)** | **1.21 (1.08,1.35)** |
| Severe constipation | 141,265 | 5,966 (4.2) | **1.27 (1.08,1.49)** | **1.78 (1.52,2.10)** | **1.26 (1.07,1.49)** |
| Epilepsy | 141,265 | 2,163 (1.5) | **1.54 (1.21,1.97)** | **1.62 (1.27,2.06)** | 1.26 (0.98,1.61) |
| Frailty | 141,265 | 9,907 (7.0) | 1.10 (0.96,1.26) | **1.61 (1.41,1.84)** | **1.28 (1.11,1.46)** |
| Alcohol misuse | 141,265 | 3,043 (2.2) | **1.39 (1.12,1.72)** | **1.41 (1.14,1.75)** | **1.55 (1.25,1.94)** |
| Peptic ulcer disease | 141,265 | 848 (0.6) | 1.46 (0.98,2.17) | **2.14 (1.43,3.19)** | **1.99 (1.32,3.01)** |

CAD coronary artery disease, CI confidence interval, HR hazard ratio, PAD peripheral artery disease, TIA transient ischemic attack
HR with 95% confidence intervals are presented.

**Figure 2.**
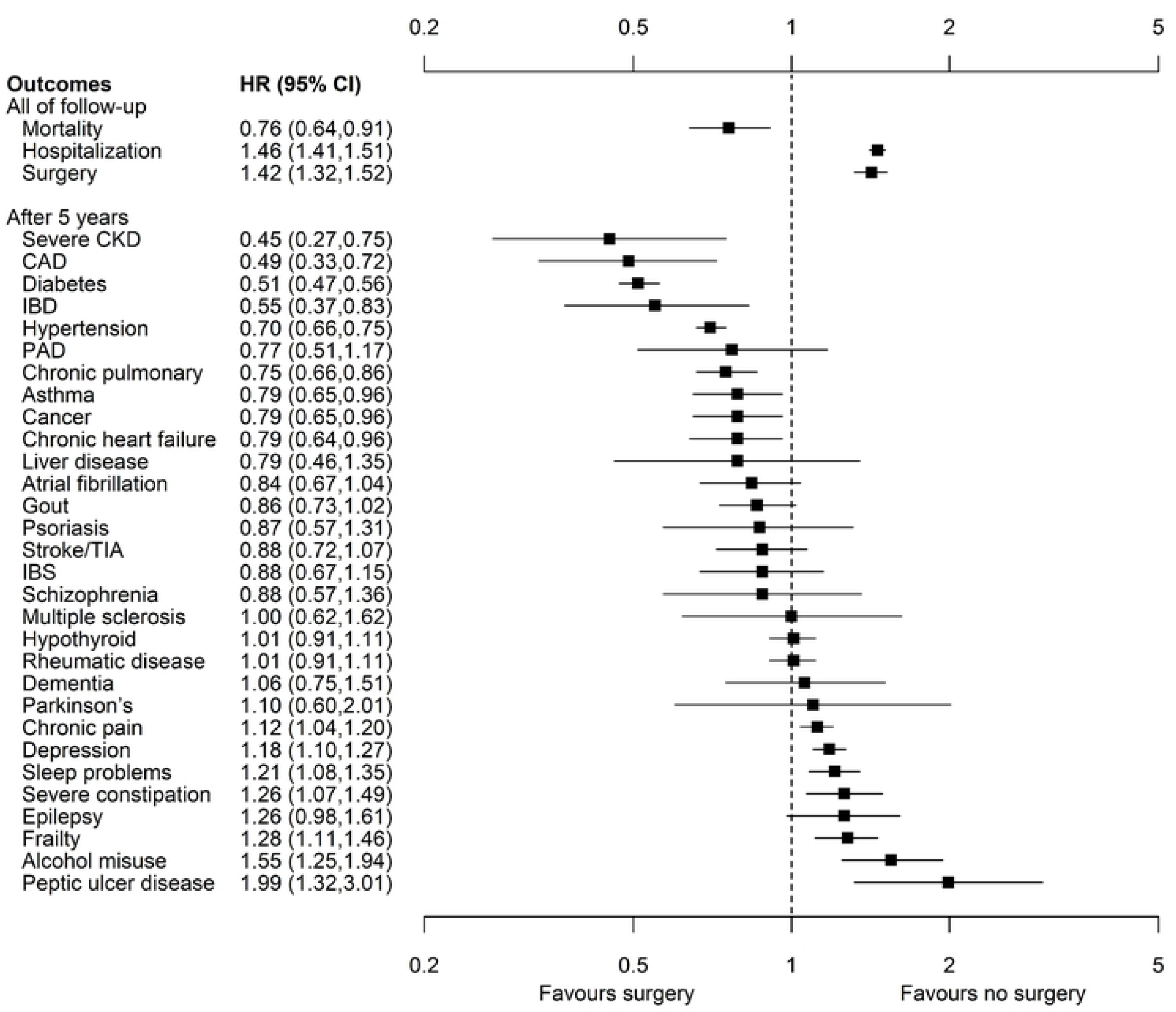
Forest plot of clinical outcomes 5-22 years after bariatric surgery. CAD coronary artery disease, CI confidence interval, CKD chronic kidney disease, HR hazards ratio, IBD inflammatory bowel disease, IBS irritable bowel syndrome, PAD peripheral artery disease, TIA transient ischemic attack

In sensitivity analyses, the fully adjusted risks for mortality in the cohort with exact matching were not significantly different for participants with and without bariatric surgery (Supplemental eTable 2). When the alternative definition of bariatric surgery was used, all results were similar to the primary analysis.

When time from the index date was parametrized with restricted cubic splines using 6 knots (Supplemental eFigure 2, top plot), the fully adjusted mortality risk associated with bariatric surgery was elevated until almost 1 year, then significantly decreased between 1 and 5 years, and then largely non-significantly decreased thereafter. Results were similar when we used 4 knots instead of 6 knots (Supplemental eFigure 2, bottom plot).

### Hospitalization and surgery

All results from unadjusted and fully adjusted models (full follow-up, HR 1.95, 95% CI 1.89,2.01; Table 2) during the first 5 years and after 5 years (Figure 2) showed that bariatric surgery was associated with excess risk of hospitalization (Table 2). Results were similar in sensitivity analyses (Supplemental eTable 2).

The hazards for time to surgery during follow-up crossed before 90 days; the magnitude of the excess risk that was associated with bariatric surgery was greater in the first 90 days but the excess risk remained throughout follow-up. Results from unadjusted, age-adjusted, and fully adjusted models (full follow-up, HR 1.42, 95% CI 1.32,1.52) demonstrated excess risk associated with bariatric surgery (Figure 2; Table 2) and were consistent in sensitivity analyses (Supplemental eTable 2). Results were similar when the number of hospitalizations or days in hospital were considered instead of time to first hospitalization, or when the number of surgeries was considered instead of time to first surgery (Supplemental eTable 3).

### Morbidities

After 5 years, bariatric surgery was associated with significantly lower adjusted risk of severe CKD (HR 0.45, 95% CI 0.27,0.75), CAD (HR 0.49, 95% CI 0.33,0.72), diabetes (HR 0.51, 95% CI 0.47,0.56), IBD (HR 0.55, 95% CI 0.37,0.83), hypertension (HR 0.70, 95% CI 0.66,0.75), chronic pulmonary disease (HR 0.75, 95% CI 0.66,0.86), asthma (HR 0.79, 95% 0.65,0.96), cancer (HR 0.79, 95% 0.65,0.96), and chronic heart failure (HR 0.79, 95% 0.64,0.96; Table 2; Figure 2).

In contrast, after 5 years, bariatric surgery was associated with significantly increased adjusted risk of PUD (HR 1.99, 95% CI 1.32,3.01), alcohol misuse (HR 1.55, 95% CI 1.25,1.94), frailty (HR 1.28, 95% 1.11,1.46), severe constipation (HR 1.26, 95% CI 1.07,1.49), sleep disturbance (HR 1.21, 95% CI 1.08,1.35), depression (HR 1.18, 95% CI 1.10,1.27), and chronic pain (HR 1.12, 95% CI 1.04,1.20).

Results were similar in sensitivity analyses using the alternative definition of bariatric surgery and when adjusting for the competing risk of death (Supplemental eTable 2). With exact matching, bariatric surgery remained significantly associated with lower risks of only diabetes and hypertension, whereas surgery remained significantly associated with higher risks of PUD, alcohol misuse, severe constipation, sleep disturbance, depression and chronic pain.

### Secular trends

The risk of mortality after bariatric surgery decreased over time from 1.13 (95% CI 0.85,1.50) in 2000 to 0.51 (95% CI 0.38,0.70) in 2015 (p=0.001; Supplemental eTable 4), as did the excess risk risk of hospitalization (from 1.63, 95% CI 1.52,1.74 in 2000, to 1.40, 95% CI 1.34,1.46 in 2015). The risk of additional surgeries did not significantly change over time.

The relative hazard of new comorbidity associated with bariatric surgery appeared stable over time for many comorbidities. However, the risks of CAD, diabetes, inflammatory bowel disease, chronic pulmonary disease and chronic heart failure, all further decreased with time (all p≤0.02) for participants with surgery versus those without, and depression and chronic pain, also, decreased with time (all p≤0.02) projecting no lower risks for participants having bariatric surgery in 2015.

### Yield of bariatric surgery

There were a total of 318 adverse outcomes averted per 400 recipients of bariatric surgery and a total of 125 adverse outcomes experienced per 400 recipients (Figure 3). The top 4 most frequent outcomes averted were diabetes, hypertension, severe CKD and CAD. The top 4 most frequent outcomes experienced were hospitalization, further surgery, depression and chronic pain.

**Figure 3.**
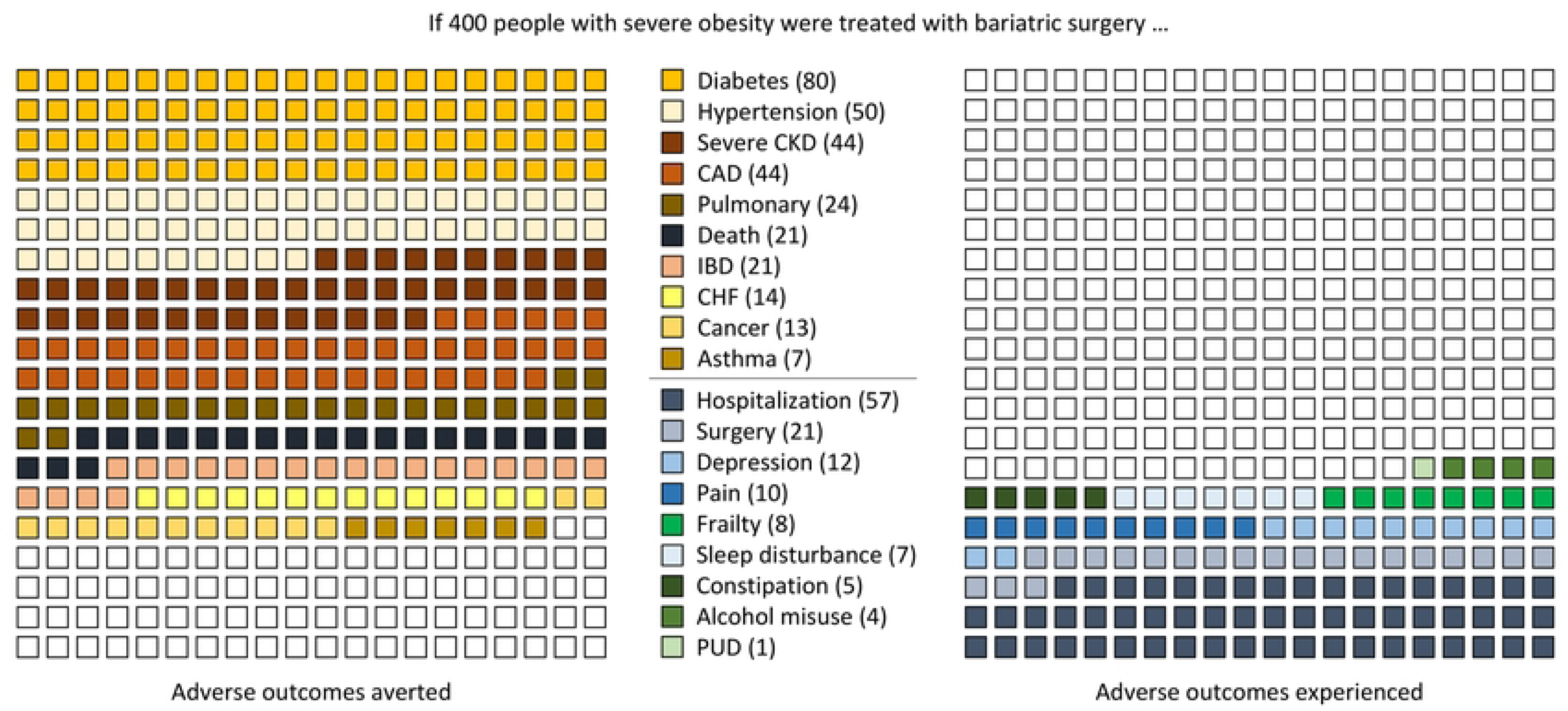
Adverse outcomes averted and experienced in 400 people. CAD coronary artery disease, CHF chronic heart failure, CKD chronic kidney disease, IBD inflammatory bowel disease, PUD peptic ulcer disease The number in the brackets in the legend adjacent to the adverse outcome gives the number of outcomes averted or experienced per 400 people. Asthma was not included in chronic pulmonary disease.

## Interpretation

Bariatric surgery was independently associated with a decreased risk of mortality (24%), and increased risks of hospitalization (46%) and surgery during follow-up (42%) (Figure 2). After 5 years, bariatric surgery was also associated with 11 to 55% lower risk of severe CKD, CAD, diabetes, IBD, hypertension, chronic pulmonary disease, asthma, cancer and chronic heart failure but with greater risk of multiple comorbidities (peptic ulcer, alcohol misuse, frailty, severe constipation, sleep disturbance, depression, and chronic pain), with excess risk ranging from 12% to 99%.

Previous studies examining the association between bariatric surgery and mortality have reported similar results. We found 5 randomized trials comparing various bariatric surgeries to standard of care, all in type 2 diabetic patients, with long-term mortality (mean/median follow-up of ≥5y) results (Supplemental eTable 5). When results were pooled using random effects meta-analysis, the risk difference was -1.5% (95% CI -4.7,1.8) and non-significantly favoured bariatric surgery. We also found 3 meta-analyses^37–39^ and a further 2 studies^40, 41^ for a total of 12 individual controlled observational studies^40–51^ exploring long-term mortality after bariatric surgery (Supplemental eTable 6). All found that bariatric surgery was associated with significantly lower mortality (range: 16% to 64% reduction), although all were potentially vulnerable to residual confounding as none adjusted for the comprehensive panel of covariates used in the current study.

Our findings for the association between bariatric surgery and the subsequent risk of morbidities extend those from previous studies. In addition to diabetes and hypertension, we found that the risks of severe CKD, CAD, IBD, chronic pulmonary disease, asthma, cancer and chronic heart failure – which are all associated with hyperinsulinemia and/or inflammation – were significantly reduced, potentially by resolving prevalent cases as well as preventing incident cases. Whether some or all of these benefits are mediated through effects on hyperinsulinemia or chronic inflammation rather than weight loss will require further investigation.

We confirmed prior work showing an association between bariatric surgery and depression, alcohol misuse and fragility fractures (a component of our frailty definition). Some studies have suggested improvements in obstructive sleep apnea with bariatric surgery.^5, 6^ We could not isolate OSA specifically in our dataset, but found an association between bariatric surgery and a higher risk of overall sleep disturbance. We further identified novel associations with an excess risk of peptic ulcer, severe constipation, and chronic pain. These adverse events all seem plausibly linked to bariatric surgery through effects on gastrointestinal structure or function (e.g., malabsorption; nutritional deficiency), although this explanation is speculative.

Our study has important strengths that increase confidence in its conclusions. First, results were consistent using two different algorithms for bariatric surgery and across a range of sensitivity analyses. Second, we studied a large population-based cohort of people drawn from a universal health care system. Third, we modelled mortality as a competing risk and again found generally consistent results to those in the main analysis.

Our study also has important limitations that should be considered when interpreting results. First, as with any observational study, residual confounding remains possible. We adjusted for at least twice as many variables as prior studies reporting on bariatric surgery and all-cause mortality: 4 demographic variables, 30 morbidities, and the earliest date of documented severe obesity. We also used a variety of sensitivity analyses and found generally consistent results. However, we could not adjust for potentially important confounders such as smoking status, BMI, fasting insulin or markers of inflammation. In order to receive bariatric surgery in Alberta, patients must be non-smokers or have ceased smoking prior to surgery,^8^ which would tend to bias our results toward the null. Additionally, since BMI is confounded by fasting insulin and c-reactive protein,^11^ lack of adjustment for BMI alone is unlikely to explain our findings. Second, those undergoing bariatric surgery were a small subset of the total cohort. Although the greater number of baseline morbidities in the bariatric group might suggest that these participants would have had worse prognosis with or without surgery, those in the control group had more serious morbidities and more hospitalizations in the 5 years prior to baseline, which suggests the opposite. Third, although we relied on administrative data rather than data on BMI to identify severe obesity, the results have face validity: data from Obesity Canada (obesitycanada.ca) found that 6.0% of Canadians had severe obesity in 2016,^52^ which is similar to the 6.1% of adult Albertans in our dataset. Fourth, while most of the ICD-9 CCP codes used in our primary administrative algorithm identified the specific type of bariatric surgery, they were not in use until later in our follow-up timeline: sleeve gastrectomy (use of 56.93C started in 2015), Roux-en-Y gastric bypass (use of 56.93A started in 2010), and adjustable gastric banding (use of 56.93B started in 2010, use of 56.93F started in 2019). Thus, we could not explore outcomes by type of bariatric surgery, since type was often unknown and also confounded potentially by any era effect.

In conclusion, bariatric surgery was associated with lower risks of certain morbidities such as diabetes, hypertension, coronary disease, severe CKD, asthma, chronic pulmonary disease, IBD, cancer and chronic heart failure as well as lower mortality. However, bariatric surgery was also associated with increased risk of hospitalization and additional surgery, as well as certain other morbidities such as alcohol misuse, osteoporosis/fragility fractures, severe constipation, peptic ulcer, sleep disturbance, depression, and chronic pain. Since values and preferences for these various benefits and harms may differ between individuals, this suggests that comprehensive counselling should be offered to patients considering bariatric surgery to help them explore this tradeoff (Figure 3), especially since not all patients will lose weight postoperatively.^15^ Future longitudinal studies should examine the effect of bariatric surgery on quality of life, as well as how quality of life may vary between patients and within patients over time.

## Data Availability

We are not able to make our dataset available to other researchers due to our contractual arrangements with the provincial health ministry (Alberta Health), who is the data custodian. Researchers may make requests to obtain a similar dataset at https://sporresources.researchalberta.ca.

## Author Contributions

NW conceived the study, wrote the first draft of the manuscript, and performed the statistical analysis. NW and MT designed the study. Both authors critically reviewed, revised, and approved the final manuscript. NW had full access to all of the data in the study and takes responsibility for the integrity of the data and the accuracy of the data analysis.

## Conflict of Interest Disclosures

None reported.

## Funding

The study was supported by MT’s David Freeze Chair in Health Services Research at the University of Calgary. The sponsors had no role in the design and conduct of the study; collection, management, analysis, and interpretation of the data; preparation, review, or approval of the manuscript; nor in the decision to submit the manuscript for publication.

## Role of Funder/Sponsor

The sponsors had no role in the design and conduct of the study; collection, management, analysis, and interpretation of the data; preparation, review, or approval of the manuscript; nor in the decision to submit the manuscript for publication.

## Disclaimer

This study is based in part by data provided by Alberta Health and Alberta Health Services. The interpretation and conclusions contained herein are those of the researchers and do not represent the views of the Government of Alberta or Alberta Health Services. Neither the Government of Alberta nor Alberta Health or Alberta Health Services express any opinion in relation to this study.

## Additional Contributions

Ben Vandermeer, MSc, University of Alberta performed a technical review; Ghenette Houston, BA, University of Alberta provided administrative support and Sophanny Tiv, BSc, University of Alberta provided graphics support.

## Notes

### Competing Interest Statement

The authors have declared no competing interest.

### Funding Statement

MT David Freeze Chair in Health Services Research at the University of Calgary https://www.ucalgary.ca/ The sponsors had no role in the design and conduct of the study collection, management, analysis, and interpretation of the data preparation, review, or approval of the manuscript nor in the decision to submit the manuscript for publication.

### Author Declarations

The institutional review boards at the Universities of Alberta (Pro00053469) and Calgary (REB16-1575) approved this study and waived the requirement for participants to provide consent due to the large sample size.

